# Identifying the drivers of multidrug-resistant *Klebsiella pneumoniae* at a European level

**DOI:** 10.1101/19012997

**Authors:** Viacheslav N. Kachalov, Huyen Nguyen, Suraj Balakrishna, Luisa Salazar-Vizcaya, Rami Sommerstein, Stefan P. Kuster, Anthony Hauser, Pia Abel zur Wiesch, Eili Klein, Roger D. Kouyos

## Abstract

Beta-lactam- and in particular carbapenem-resistant Enterobacteriaceae represent a major public health threat. Despite strong variation of resistance across geographical settings, there is limited understanding of the underlying drivers.

To assess these drivers, we developed a transmission model of cephalosporin- and carbapenem-resistant *Klebsiella pneumoniae*. The model is parameterized using antibiotic consumption and demographic data from eleven European countries and fitted to the resistance rates for *Klebsiella pneumoniae* for these settings. The impact of potential drivers of resistance is then assessed in counterfactual analyses.

Based on reported consumption data, the model could simultaneously fit the prevalence of extended-spectrum beta-lactamase-producing and carbapenem-resistant *Klebsiella pneumoniae* (ESBL and CRK) across eleven European countries over eleven years. The fit could explain the large between-country variability of resistance in terms of consumption patterns and fitted differences in hospital transmission rates. Based on this fit, a counterfactual analysis found that reducing nosocomial transmission and antibiotic consumption in the hospital had the strongest impact on ESBL and CRK prevalence. Antibiotic consumption in the community also affected ESBL prevalence but its relative impact was weaker than nosocomial consumption. Finally, we used the model to estimate a moderate fitness cost of CRK and ESBL at the population level.

This work highlights the disproportionate role of antibiotic consumption in the hospital and of nosocomial transmission for resistance in gram-negative bacteria at a European level. This indicates that infection control and antibiotic stewardship measures should play a major role in limiting resistance even at the national or regional level.

**Significance Statement:** As beta-lactam resistant gram-negative bacteria represent one of the most critical threats in the ongoing antibiotic resistance crisis, it is crucial to identify the underlying drivers and appropriate measures to curb their spread. By combining a transmission model with epidemiological data at a European level, we can explain the strong differences of extended-spectrum beta-lactamase-producing and carbapenem-resistant *Klebsiella pneumonia* across countries and their often-rapid temporal increase. We find that among potentially modifiable drivers, inpatient antibiotic consumption and nosocomial transmission rates have the strongest impact on resistance. This implies that infection control and antibiotic stewardship in hospitals are crucial for preventing antibiotic resistance in gram-negatives even beyond individual hospitals as they may affect resistance prevalence at the level of entire countries.

## Introduction

Beta-lactam resistant *Enterobacteriaceae* represent one of the most serious threats in the current antimicrobial resistance crisis (1). Accordingly, carbapenem-resistant and extended spectrum beta-lactamase (ESBL) producing *Enterobacteriaceae* are examples mentioned by the World Health Organization (WHO) in the last “Prioritization of Pathogens to Guide Discovery” (1). Beta-lactams are a widely used type of antibiotics due to their broad spectrum of activity against Gram-negative bacteria. *Enterobacteriaceae* are common commensal flora, particularly in the gastrointestinal tract, and are typically exposed to any antibiotic treatment administered to an individual. Hence, they have developed resistance to most of the commonly used antibiotics. For example, carbapenem-resistant organisms (CRO) are resistant to all known beta-lactams (2). Last-resort drugs, such as colistin, generally remain effective, though there have already been reported cases of *Enterobacteriaceae* resistant to both carbapenems and colistin (3). While newer drugs, such as ceftazidime-avibactam, have been introduced, widespread dissemination of carbapenem resistant genes may herald the beginning of a post-antibiotic era (4), at least for the Enterobacteriaceae species in question.

*Klebsiella pneumoniae* is one of the leading causes of hospital-acquired infection (HAI), and 5-30% of the general population are colonized with it (5). In general, *K. pneumoniae* behaves as commensal flora. Thus, exposure to antibiotics remains undetected, as antibiotic prescription is rarely associated with infections. There are several studies that have found a significant correlation between antibiotic use and the prevalence of antibiotic resistance in a variety of pathogens (6–10). However, these correlations are usually far from perfect, e.g. higher consumption does not always indicate more resistance when comparing countries, indicating that other drivers may be at least as important as antibiotic consumption in determining levels of antibiotic resistance(11).

In order to optimize prevention measures, it is critical to identify the drivers of resistance and to understand how interventions targeting those drivers would translate into changes in antibiotic resistance. Antimicrobial resistance is affected by a number of potential drivers such as the consumption of antibiotics in the human population, consumption in livestock, health care-related transmission, travel, and environmental contamination (11).One of the most notable drivers of resistance is overall antibiotic consumption in humans (6–10).

However, antibiotic consumption in humans is not uniformly distributed, but is rather characterized by strong heterogeneities imposed by demographics and the institutional setting (11). One particularly relevant instance of this structure of antibiotic consumption are the differences between the hospital and community settings, as per-capita consumption and transmission rates tend to be higher in the hospital setting (5). The effects of population structure (ranging from travel, to livestock, to institutional setting) are in principle detectable by genomic and molecular epidemiology approaches (12–14). However, while such approaches can help to characterize individual outbreaks, the high frequency of asymptomatically colonized individuals and the fact that these individuals are typically not sampled, implies that it is extremely challenging to obtain a representative assessment of the overall importance of different settings with these methods. In this context, computational models offer a unique opportunity to understand how antibiotic consumption, its distribution by setting, and the transmission of pathogens in hospitals contribute to antibiotic resistance at the population level.

Here, we aim to combine epidemiological models for the spread of resistance with surveillance data on antibiotic consumption and resistance, in order to determine the key driving factors of the spread of CROs and ESBL. We focused on rates of resistance for *K. pneumoniae*, as it is one of the most common causes of bloodstream infections and hospital-acquired pneumonia (15), and mortality rates related to infection are high. The mortality of bloodstream infection caused by carbapenem-resistant *K. pneumoniae* (CRK) has been estimated to be 50% (16).In addition, the epidemiology of this pathogen is well monitored by the European Center for Disease Prevention and Control (ECDC) for several countries (17),and resistance rates are highly variable across countries, with some countries exhibiting low levels of 3^rd^ generation cephalosporin resistance and almost no carbapenem resistance (Denmark, Finland, Netherlands, Norway, Sweden), whereas in other countries carbapenem resistance levels are above 20% (e.g. Greece, Italy). Antibiotic resistant *K. pneumoniae* provides thus a good case study for understanding the variability and drivers of antibiotic resistance.

## Methods

### Model

We used a deterministic compartmental model to simulate the spread of ESBL and CRK in the hospital and the community. Our model has three principal dimensions: setting, colonization, and treatment (see Figure 1 B): we stratified the population into hospital and community settings to represent the difference in antibiotic consumption and transmission between the two settings. All individuals were classified by colonization status into susceptible, colonized (i.e. asymptomatic carriers of *K. pneumonia)*, or infected (i.e. with symptoms caused by *K. pneumoniae)*. Colonized and infected individuals were also stratified by strain as non-resistant, ESBL, and CRK. The susceptible and colonized compartments could either be treated with 3^rd^/4^th^ generation cephalosporins (drug A) or carbapenems (drug B) or not treated at all. As most *K. pneumoniae*-colonized individuals who are exposed to antibiotics are treated for unrelated illnesses, we assumed that this treatment is not affected by colonization status and strain.

**Figure 1.**
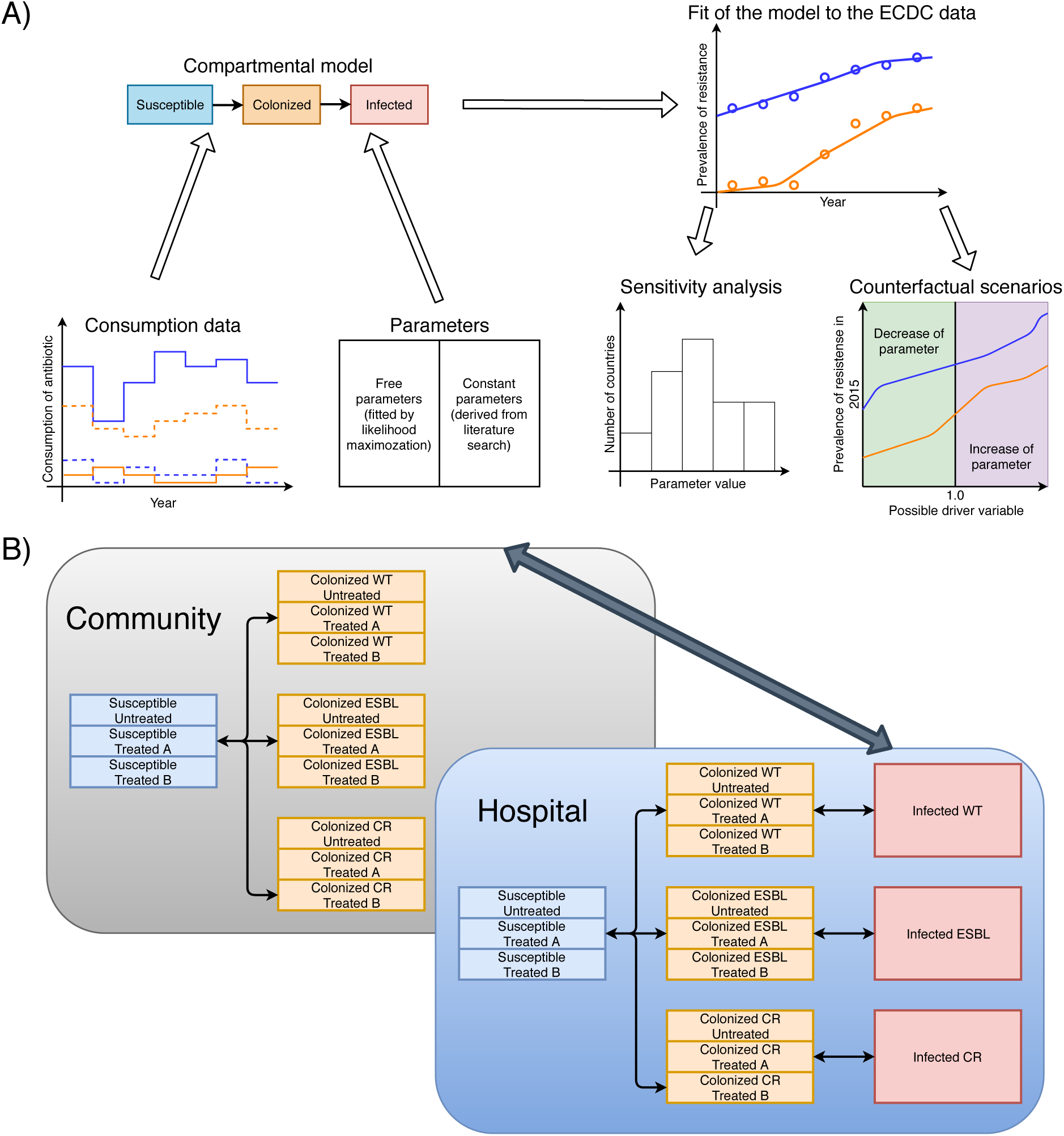
The workflow of the modeling approach. Consumption and resistance data were acquired from ECDC, and other parameters were found in the literature or used as free parameters. The model was fit to the data reported by ECDC to optimize the free parameters. Sensitivity analyses were performed to test the robustness of the model. Counterfactual scenarios were applied to understand the functional dependencies of the prevalence of resistance from possible drivers.

All infected individuals were assumed to be in the hospital setting. If an infection occurs in the community, the individual was assumed to be hospitalized immediately upon the development of symptoms. We further assumed that symptomatically infected individuals are properly diagnosed and appropriately treated. This may be too optimistic, but it should be noted that the number of symptomatically infected individuals is small (compared to the colonized individuals) and hence their contribution to both consumption and transmission of resistance is negligible. We introduced the symptomatically infected compartments to model the sampling process, not for measuring their influence on consumption and transmission (which is negligible). This was done, because all reported samples in ECDC data were collected for bloodstream infections and spinal fluid infections.

Colonization can occur due to contact with colonized individuals and due to import from external sources (which may reflect any process not explicitly captured in the model, for example travel, agriculture etc.). In addition, we assumed resistance can spread due to super-colonization followed by horizontal gene transfer. By this process individuals colonized with a sensitive strain can acquire resistance (this rate is however lower than primary colonization, see Tables 1, S1, and S2 and the term HGT in the supplementary material equations). To include import of colonized strains from other countries and from agriculture, we added a constant extrinsic force of colonization as a free parameter to our model. Decolonization can happen due to the treatment by antibiotics or due to natural clearance rates. The complete description of the processes in the model and the model equations are available in the Supplementary Material.

**Table 1:** Free model parameters of the fit

| Parameter | Variable hospital transmission rates across countries | Same hospital transmission rate for all countries |
| --- | --- | --- |
| Fitness cost ESBL | 1.97% | 1.34% |
| Fitness cost CRK | 2.26% | 1.34% |
| Import of ESBL (reservoir size*) | 327.1 per 100000 persons | 80.6 per 100000 persons |
| Import of CRK (reservoir size*) | 4.6 per 100000 persons | 0.35 per 100000 persons |
| Colonization rate | $9.8 \cdot 10^{-3} \text{ day}^{-1}$ | $1.0 \cdot 10^{-2} \text{ day}^{-1}$ |
| Super-colonization coefficient | 0.186 | $1.21 \cdot 10^{-6}$ |
| Increased susceptibility by treatment | 0.11 | 0.06 |
| Displacement/loss of plasmid rate (relation to natural decolonization rate) | 0.046 | $2.2 \cdot 10^{-4}$ |
|  | Hospital transmission rate (relative to the community level) |  |
| Greece | 31.7 | 29.2 |
| Italy | 20.5 |  |
| Portugal | 18.5 |  |
| Croatia | 13.9 |  |
| France | 11.3 |  |
| Hungary | 12.1 |  |
| Denmark | 14.0 |  |
| Finland | 0.44 |  |
| Netherlands | 7.7 |  |
| Norway | 8.8 |  |
| Sweden | 10.7 |  |
| Log-likelihood | -1001 | -1226 |
| BIC | 2220 | 2616 |
| p-LRT | <0.001 |  |

\* Import of resistance strains is included as a constant term added to the force of infection. For interpretability and given the form of the force of infection (see supplementary material section 1.4), this term is expressed here as the equivalent of the force of infection that would have been caused by a given number of individuals colonized by the resistant strain.

### Fitting process

To calibrate the model and determine the free parameters such as fitness costs of resistance, we fitted the model to the resistance data reported by the ECDC. This fit was done by maximization of likelihood, assuming a binomially distribution of data (see supplementary material section 2.1). We compared two main scenarios: in the first case, we assumed that each country had a unique nosocomial transmission rate, while in the second case all countries were assumed to have the same transmission rate. Based on these fits, we assessed possible drivers of resistance by using counterfactual scenarios. Specifically, we considered four potential drivers: nosocomial transmission rate, inpatient and outpatient consumption of 3^rd^ and 4^th^ generations cephalosporins, and inpatient consumption of carbapenems. Finally, to evaluate the robustness of our results, we performed two types of sensitivity analyses: firstly, a leave-one-out analysis, where we excluded one of the countries and fitted the model to the remaining ten countries; and, secondly, variation of fixed parameters with high uncertainty (colonization prevalence, time of treatment, time of clearance on treatment, mean length of colonization, time of disease development in hospital) in a multivariable sensitivity analysis.

### Data

We parametrized and calibrated the model using different types of data (see supplementary material and in particular supplementary table S1, S2): consumption, hospitalization rate, and length of hospitalization, which were obtained from surveillance data from the ECDC and WHO. The biological and epidemiological parameters for which estimates were available were extracted from the literature (supplementary table S1, S2).

We retrieved annual data on the prevalence of resistant bacterial strains and antibiotic consumption from the ECDC. Data on resistance was collected through the European Surveillance System (TESSy) by the ECDC, which includes data going back to 2005 for 30 countries (17). Consumption data covers the same time range and includes both hospital and community consumption rates (18). Countries were included if they had both sufficiently complete data for resistance to 3^rd^ generation cephalosporins and carbapenems and for use of 3^rd^ and 4^th^ generations cephalosporins and carbapenems from 2005 to 2015 (see flowchart on Figure S4).

We assume resistance to 3^rd^ generation to be a good proxy for the ESBL strain. In line with ECDC reports, considering the fact that between 65.2% and 100% of 3^rd^ generation cephalosporin isolates are ESBL-positive, we assumed resistance to 3^rd^ generation cephalosporins to be a proxy for ESBL strains (19). We excluded countries that had less than six out of ten annual records for antibiotic consumption in the hospital setting. Furthermore, we excluded countries that had less than 18 resistance entries out of the 22 possible. Also, for 4 of them there are less than 18 resistance entries with the number of reported samples being more than 200. As a result, we restricted our analysis to 11 countries (Croatia, Denmark, Finland, France, Greece, Hungary, Italy, Netherlands, Norway, Portugal, Sweden) with sufficient data on both consumption and resistance (Figure S4).

## Results

We combined a mathematical model for the transmission of ESBL and CRK with antibiotic resistance and consumption data for 11 European countries in order to investigate the drivers of resistance evolution (see Figure 1 and Methods and Supplementary material).

Qualitatively, the European countries considered here can be divided into three main groups (Figure 3). The first group consists of countries with high prevalence of resistance to both 3^rd^ generation cephalosporins and carbapenems (Greece and Italy) (prevalence of carbapenem resistance higher than 30% and prevalence of resistance to 3^rd^ generation cephalosporins higher than 50%). The second group consists of countries with high prevalence of resistance to 3^rd^ generation cephalosporins but low prevalence of resistance to carbapenems (Croatia, France, Hungary, Portugal) (prevalence of carbapenem resistance less than 10% and prevalence of resistance to 3^rd^ generation cephalosporins higher than 30%). Finally, the third group consists of countries with low prevalence of resistance to both (Denmark, Finland, Netherlands, Norway, Sweden) (prevalence of carbapenem resistance less than 3% and prevalence of resistance to 3^rd^ generation cephalosporins less than 15%).

The correlation between the total (inpatient plus outpatient) consumption of 3rd and 4th generation cephalosporins and the prevalence of resistance is weak (adjusted R^2^=0.27). However, the corresponding correlation with inpatient consumption is stronger (R^2^=0.51) (see Figure 2 A). As the consumption of carbapenems selects for the resistance to both carbapenems and cephalosporins (because CRK are also resistant to cephalosporins), it is reasonable to consider both 3^rd^ and 4^th^ generation cephalosporins and carbapenems as drivers for the spread of resistance to 3^rd^ generation cephalosporins. Indeed, in this case the correlation is even higher (R^2^=0.64). Finally, the strength of the correlation with consumption rates can change considerably if the average yearly change of resistance is considered instead of the prevalence of resistance (Figure S5). These different correlations provide a first indication that the structure of antibiotic use (inpatient vs. outpatient), the consumption of other antibiotics in the same class, and the dynamics of resistance should be taken into account for understanding the association between antibiotic use and resistance. For carbapenem resistance, the association between consumption and resistance prevalence is even weaker (Figure 2 B). For example, both the Italian and Greek levels of carbapenem consumption are comparable with other countries (Portugal, Hungary, Finland) which do not exhibit a strong increase in CRK.

**Figure 2.**
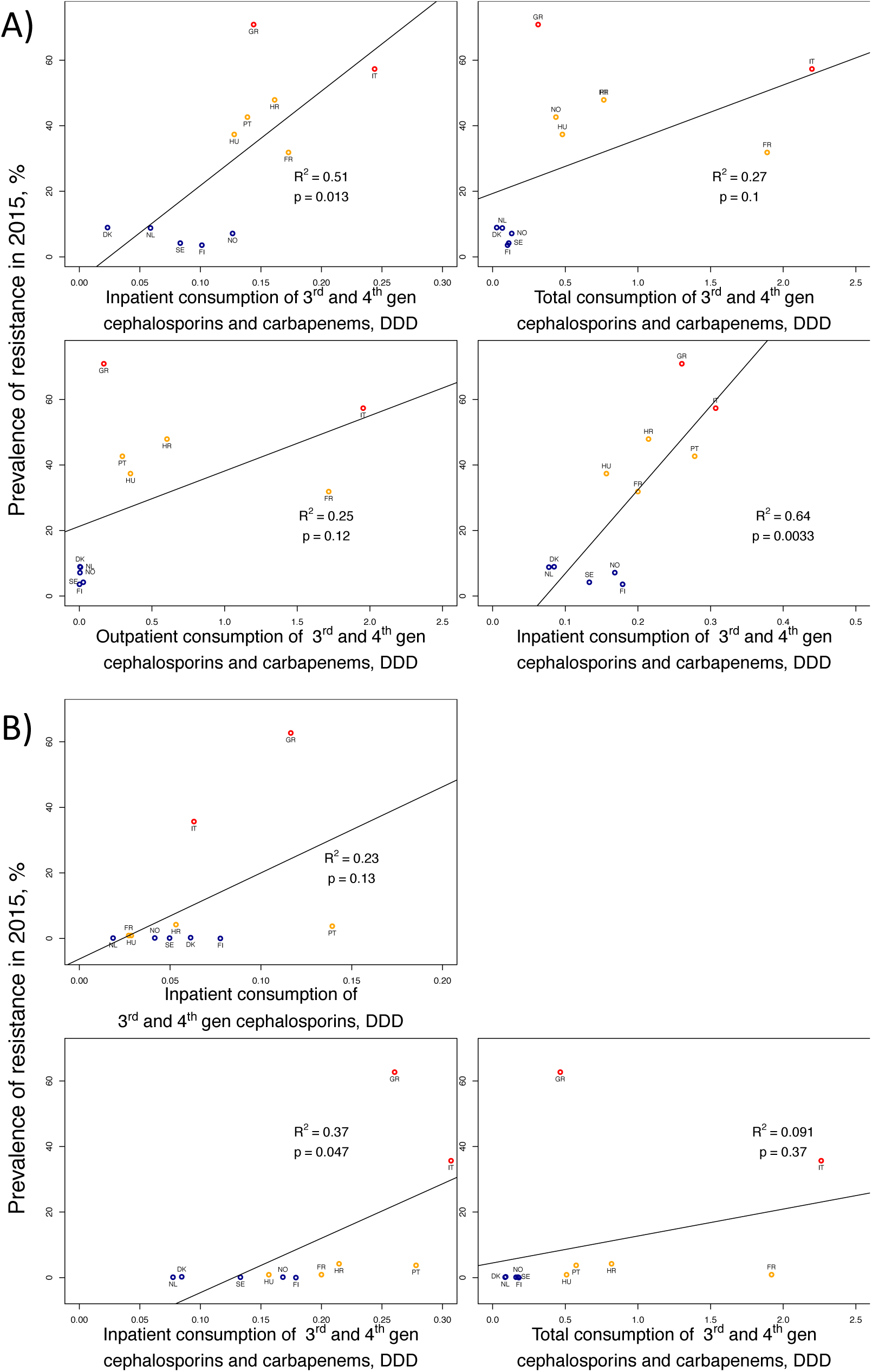
Correlation between the consumption of different classes of antibiotics in different settings (x-axes), and the prevalence of resistance to 3rd generation cephalosporins (a), prevalence of resistance to carbapenems (b).

The eleven included countries exhibited qualitatively different time courses of resistance and consumption (Figures S1-S3). We fit our model by varying among the free parameters only hospital transmission rate across countries and keeping the other free parameters constant across countries (see Table 1). This corresponds to the assumption that biological parameters are comparable across countries, while transmission in the hospital, which depends on nosocomial infection prevention, is setting specific. The model fit shows a considerable variation in hospital transmission rates, which range from 1 times to 30 times the corresponding rate in the community (Table 1). Overall, we find that this model can capture both the dynamics within and the variability across the eleven European countries considered (Figure 3).

**Figure 3.**
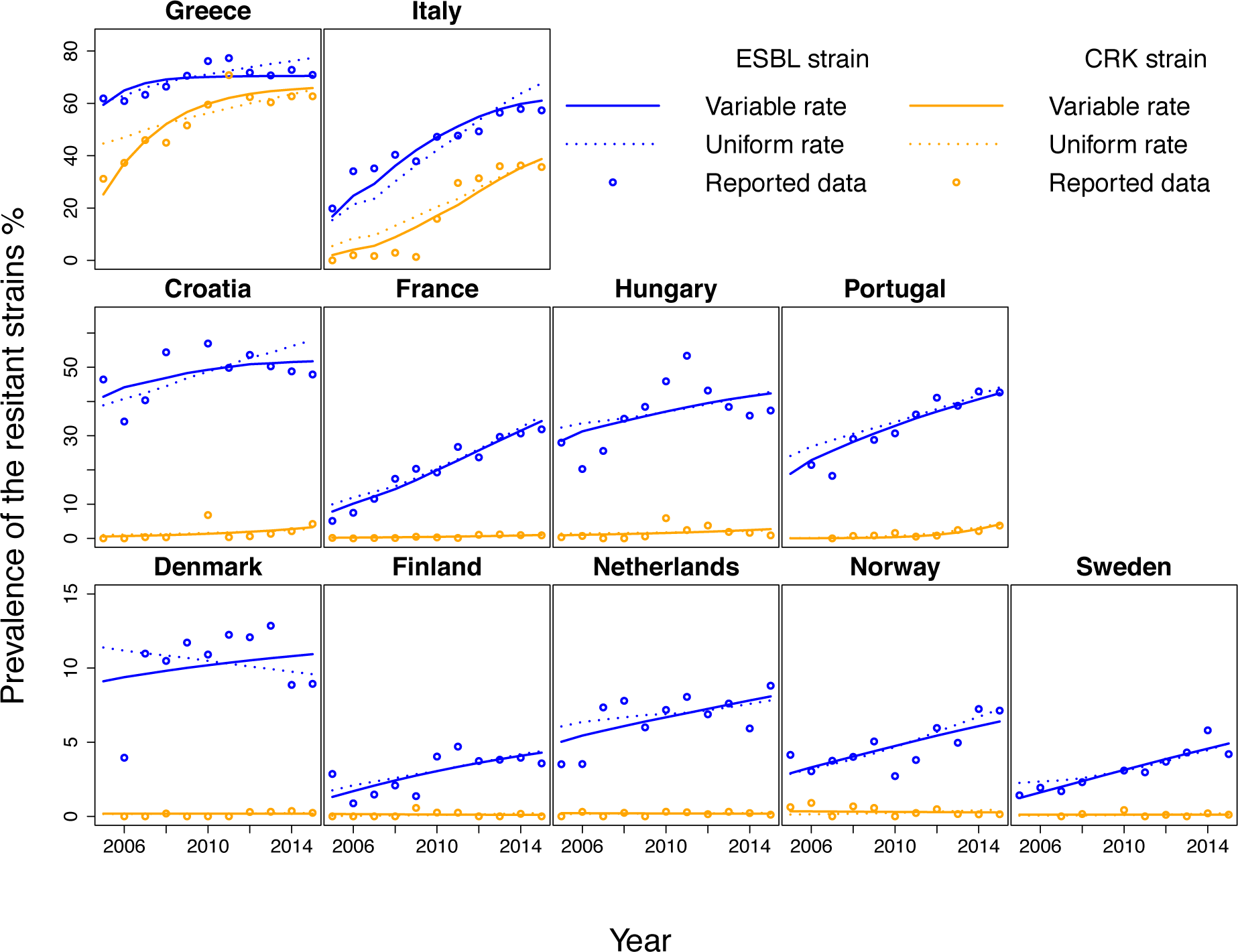
Model fit of ESBL and CRK. The model was fitted to the data of the annual prevalence of resistance in *Klebsiella pneumoniae* reported by ECDC from 2005 to 2015. Circles represent the reported data and solid and dotted lines represent the fit with variable between-country and uniform for all hospital transmission rates respectively.

We find that assuming one universal hospital transmission for all countries provides a significantly worse fit of antibiotic resistance levels than the model allowing this rate to vary across countries (Figure 3 and Table 1). Even though the model with a universal transmission rate provides overall a qualitatively acceptable fit for most countries, it misses several important features of the dynamics of resistance in the individual countries. Firstly, the model fails to reproduce some of the extreme cases among very high and low prevalence countries. For example, it could not capture the emergence of carbapenem resistance in Italy in 2010-2011 from near zero levels to over 30%, or the slight decrease of carbapenem resistance in Norway (see Figure 3. Secondly, the fitted initial levels of resistance strongly differ in this model for many countries (Greece, Italy Portugal) from the ECDC data, which again reflects the model’s inability to capture extreme changes in antibiotic resistance.

By applying counterfactual scenarios, we found that nosocomial transmission and the structure of antibiotic consumption played a key role as drivers of both carbapenem but also ESBL strains. To determine the role of nosocomial transmission for the spread of ESBL and CRK, we varied the corresponding inpatient transmission rate over a broad range (Figure 4). We found that hospital transmission affected the level of resistance to carbapenems and also the prevalence of ESBL strains (Figure 4). Despite this, in some countries such as Finland and Norway, hospital transmission plays a minor role because it is low overall (see Figure 4 and Table 1). Nevertheless, the results indicate that hospital transmission is a major driver of the spread of both ESBL and carbapenem-resistant *K. pneumoniae* strains. Concerning the effect of the structure of antibiotic consumption, we found that antibiotic use in both the hospital and community setting affects resistance, but that consumption in the hospital has a stronger effect: even for ESBL, relative changes of the consumption of cephalosporins in hospitals has overall a slightly stronger impact than of the outpatient consumption (Figure 5), despite the fact that the absolute amount of 3^rd^ and 4^th^ generation cephalosporins consumed in the community is considerably higher than that in the hospital (Figure S1 S2). This implies that the effect of a given absolute amount of antibiotics (e.g. a given number of DDDs) is larger if it is consumed in the hospital than if is consumed in the community. Our results also show that carbapenem consumption could be a selective factor for the resistance to 3rd generation cephalosporins (Figure 5) and that high consumption levels of 3rd generation cephalosporins can affect the level of carbapenem resistance (Figure 5, for Italy). In addition, import of resistance from other countries and agriculture could play a key role in the spread of ESBL-strains in low-prevalent countries (Figure 6), despite the fact that the import rate is low. Finally, we find also in the model assuming a uniform nosocomial transmission rate across countries that transmission and consumption in hospitals are key drivers of resistance and that import is mainly of importance for low-prevalence countries (Figures S6-S8).

**Figure 4.**
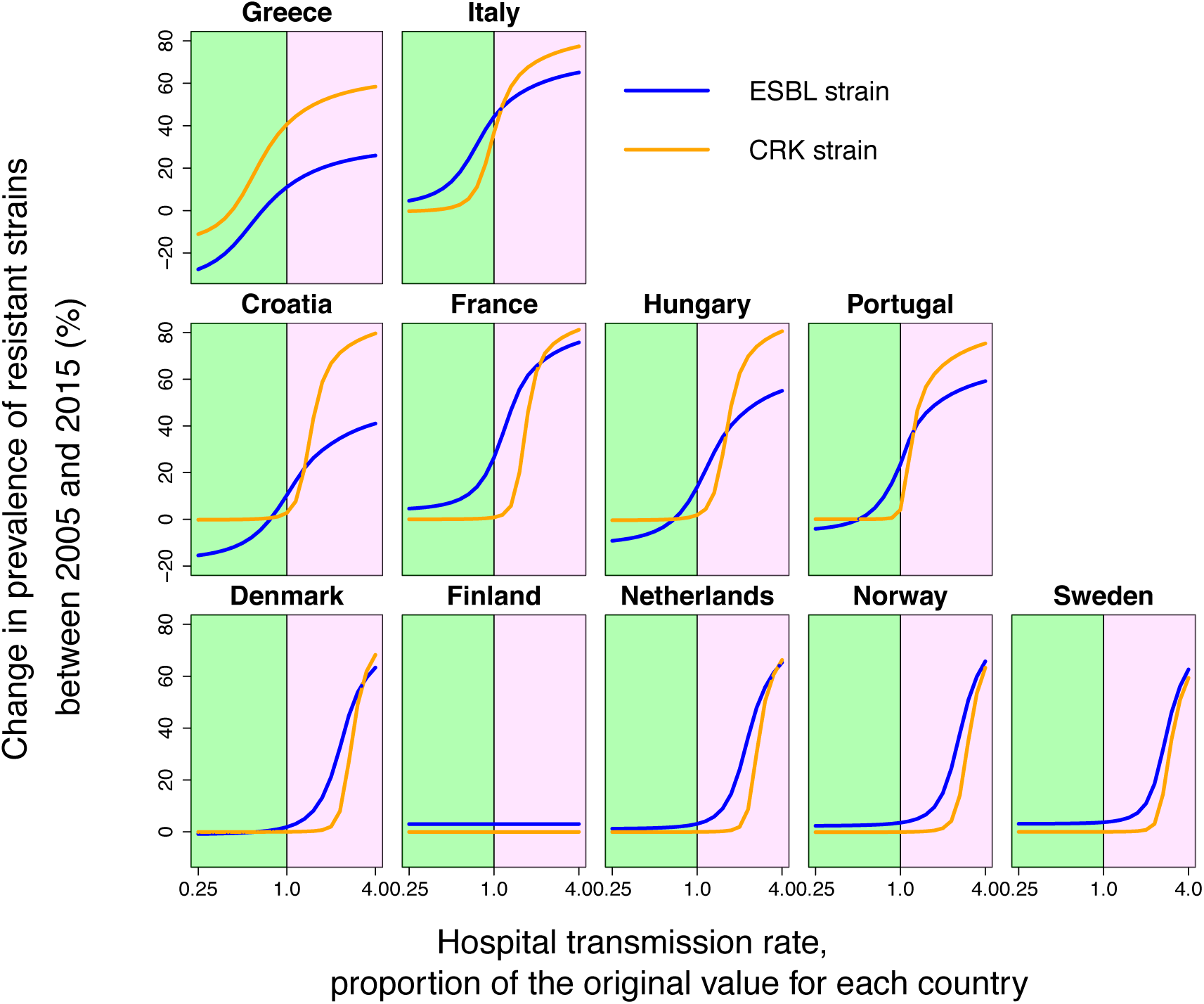
Counterfactual scenarios observed by the variation in hospital transmission rate (from 0.25 to 4.0 of the original value). Plots represent the dependence of change in prevalence of resistant strains between 2005 and 2015 from the level of the hospital transmission rate. Green and purple areas represent the decrease and increase in hospital transmission rate respectively.

**Figure 5.**
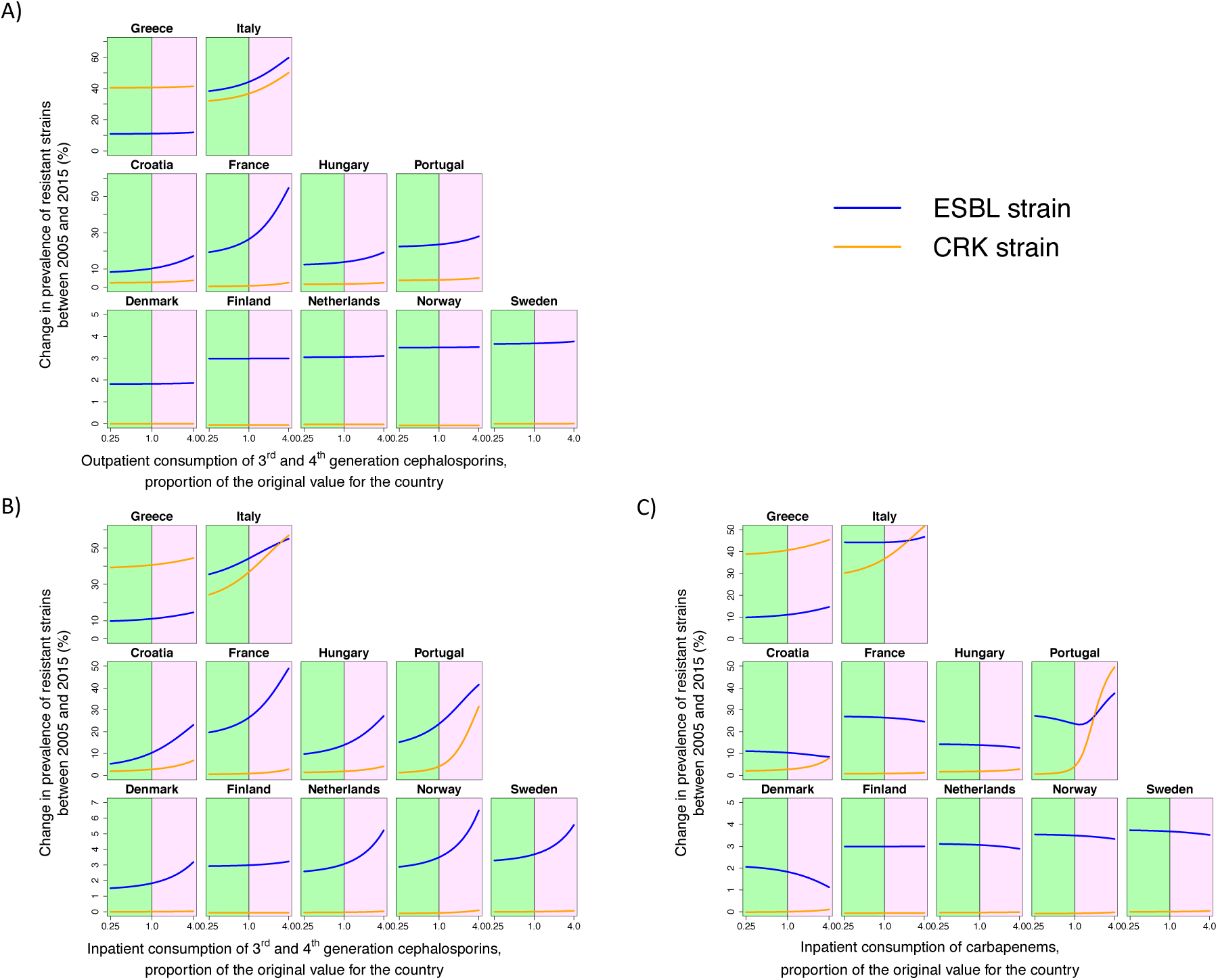
Counterfactual scenarios observed by the variation in consumption of different antibiotic classes (from 0.25 to 4.0 of the original value). Plots represent the dependence of change in prevalence of resistant strains between 2005 and 2015 from the level of antibiotic consumption. Green and purple areas represent the decrease and increase in hospital transmission rate respectively. A) Outpatient consumption of 3^rd^ and 4^th^ generation cephalosporins B) Inpatient consumption of 3^rd^ and 4^th^ generation cephalosporins C) Inpatient consumption of carbapenems

**Figure 6.**
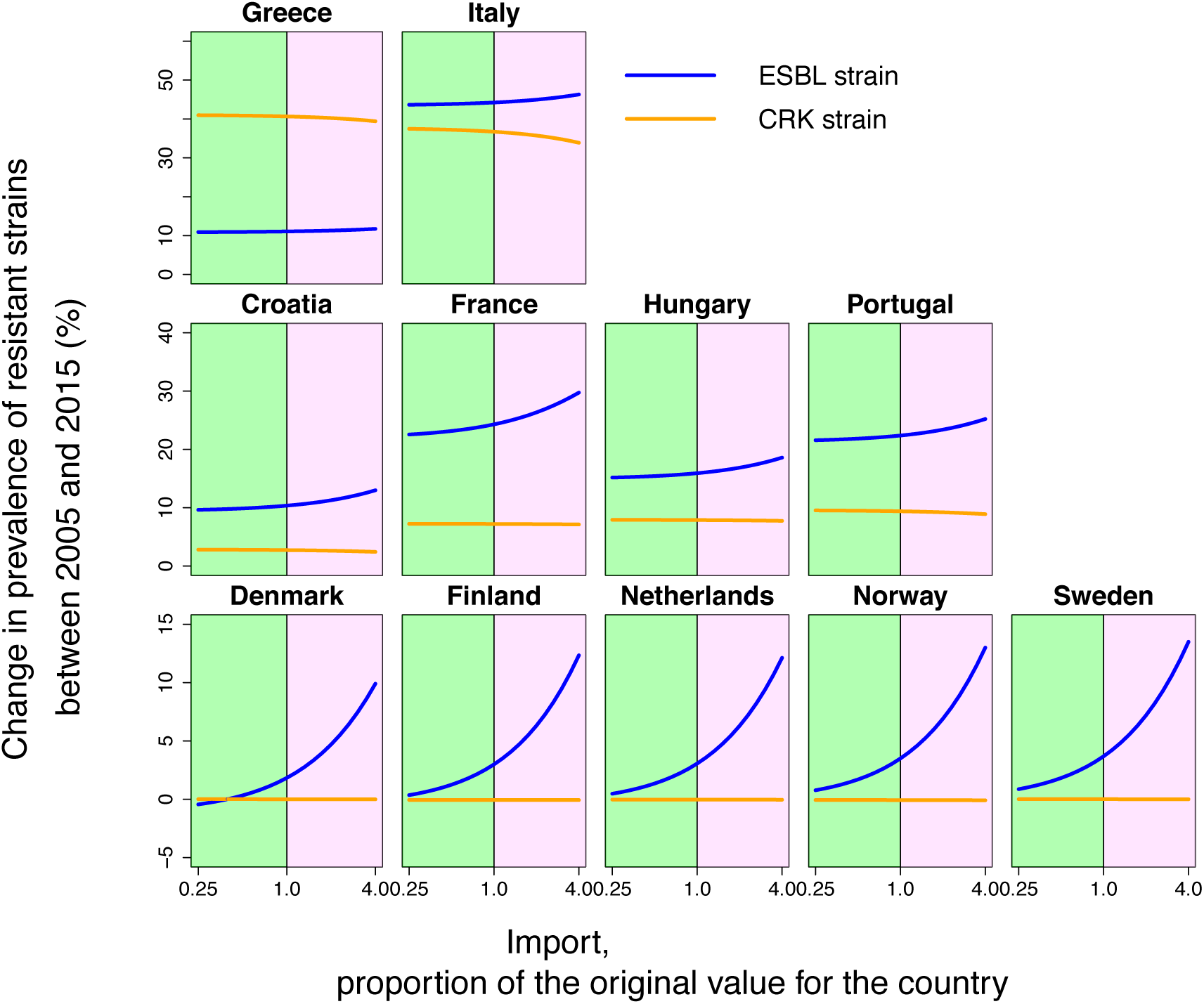
Counterfactual scenarios observed by the variation in import of ESBL strain (from 0.25 to 4.0 of the original value). Plots represent the dependence of change in prevalence of resistant strains between 2005 and 2015 from the level of the hospital transmission rate. Green and purple areas represent the decrease and increase in hospital transmission rate respectively.

We performed two types of sensitivity analyses to assess the robustness of our results. Firstly, a leave-one-out analysis assessed the dependence of our estimates on the inclusion of individual countries (Figures S9 and S10). Secondly, we have performed a multivariate sensitivity analysis simultaneously varying 5 parameters (colonization prevalence, time of treatment, time of clearance on treatment, mean length of colonization, time of disease development in hospital) within the ranges indicated in Table S2 (Figure S11). Overall, these analyses showed that the above results were robust to both variation of fixed parameters and removal of individual countries from the analyzed data set.

## Discussion

The epidemic model presented here could explain the spread of carbapenem resistance and ESBL strains over eleven years for eleven European countries with diverse resistance rates and trajectories. In particular, we found that a good fit of the observed resistance data was possible while only varying the hospital transmission rate but keeping the rest of the parameters constant across countries. The model fit provided estimates of key unknown parameters, in particular the fitness cost associated with antimicrobial resistance. Using counterfactual scenarios, our results suggest that the hospital environment, both in terms of transmission and antibiotic consumption, plays a key role for the spread of antimicrobial resistance even at the level of entire countries.

Previous studies have shown for several pathogen-drug combinations significant correlations between antibiotic consumption and the prevalence of resistance (6, 9, 20–22). However, European data for *K. pneumoniae* exhibit no simple relationship between levels of consumption and resistance. Using a dynamical modelling approach to link the history of consumption and resistance allowed us to explain these apparent discrepancies, and to provide a mechanistic explanation for the difference across countries and for the rapid dynamics of resistance. In particular, we found two factors to be central: the structure of antibiotic consumption (hospital vs. community) and nosocomial transmission of *K. pneumoniae*:

Even though overall the majority of beta-lactams are consumed in the community, we found that inpatient consumption may be a critical factor for the spread of resistance.

Specifically, our results indicate that a relative change of 3^rd^ generation cephalosporins consumption in the hospital has a similar or even higher impact than the same relative change in the community (Figure 4). The absolute amount (in DDDs) of 3^rd^ generation Cephalosporins consumed in the hospital is however considerably lower than in the community (Figure S1, S2). This implies that an absolute change in antibiotic consumption (e.g. by a given number of DDDs) has a much higher impact if it occurs in the hospital than if it occurs in the community. Intuitively, this can be explained by the fact that despite absolute levels of antibiotic consumption being lower in the hospital, the relative consumption per patient-time is higher than in the community (in terms of DDD per person-time). Thus, the hospital setting can act as an environment where resistant strains have a selective benefit, leading to a source-sink constellation (23) with the hospital representing the source and the community the sink for resistance. Moreover, due to its higher transmission rate, the hospital can turn into a hotspot of colonization with the resistant strain (especially in the high-prevalence countries), explaining the disproportionate impact of antibiotic consumption we observed in the counterfactual scenarios, where even for 3^rd^/4^th^ generation cephalosporins, consumption in the hospital had a much stronger impact on the corresponding resistance evolution than consumption in the community. As a consequence, our findings also imply that overall levels of antibiotic consumption may not be the optimal way to summarize the impact of consumption on resistance. Instead, a DDD consumed in a high-transmission setting may have a much stronger impact than a DDD consumed in a low-transmission setting, implying that consumption rates should ideally be weighted or stratified by the environment they are consumed in.

Similar to antibiotic consumption in the hospital, we found that nosocomial colonization rates play an important role both in terms of the counterfactual scenarios considered and in terms of explaining differences between countries. Again, this is consistent with the notion of the hospital environment representing a hotspot for the transmission of antimicrobial resistance even against drugs that are primarily consumed in the community. The high variability of hospital transmission/colonization rates observed between countries can thus explain why countries with similar levels of consumption exhibit different levels of resistance. In turn, this variability of estimated transmission rates is expected to be affected by a range of factors such as investment in hospital hygiene and infection control or hospital occupancy and distribution patterns (number of patients per room, structure within hospitals etc.).

Our results suggest thus that both consumption and transmission rates in the hospital are critical drivers for the spread of resistance (24). This indicates that investments in infection control may not only benefit the individual hospital making those investments but can also have an impact on the level of resistance at the country level. In line with (25), we found that such collateral benefits are strongly dependent on the epidemiological setting. Hence, the possibility of such collateral benefits are consistent with the success of several public health interventions to reduce transmission in hospitals (26).The impact of the structure of the consumption suggests that measures which would shift hospital consumption of antibiotics to the community would give a benefit in terms of slowing down the spread of resistance (such a shift could for example be achieved by outpatient intravenous antibiotic treatment).

Moreover, our results suggest that resistance to a particular antibiotic could depend on the consumption of other antibiotics of the same class.

Considering the qualitative behavior of our model across countries, we found three main types of possible settings: Firstly, countries with high prevalence of resistance and high hospital transmission rates, which plays a dominant role in the spread of resistance. It is notable that in some of these countries (in particular in Greece), hospital transmission rates were estimated to be so high that the model predicts the spread of resistance to be almost independent from antibiotic consumption rates. Secondly countries with medium prevalence, where the spread is mostly driven by the antibiotic consumption and especially the antibiotic consumption in hospitals. Thirdly, countries with low prevalence characterized by low hospital transmission rates, where import of resistance is a key factor.

Finally, our model could also be used to estimate unknown parameters governing the spread of resistance. One such key parameter determining the spread of resistance is the fitness cost that resistant strains pay in the absence of antibiotic treatment. However, such fitness costs are notoriously difficult to estimate. While it is possible to measure competitive differences *in vitro*, the relevance of such measures for strain competition at the epidemiological level is uncertain. The modelling approach presented here offers a possibility to obtain such fitness cost estimates from the model fit. Intuitively, these estimates are the parameter values of the fitness cost for which the observed levels of consumption would lead to the observed levels of resistance. The estimated values (Table 1) indicated weak but non-negligible fitness costs, which is consistent with *in vitro* estimates (27, 28).

Our model has several limitations and strengths. Like every model it is based on simplifying assumptions which are mainly dictated by the (granular) availability of data and the difficulties of parametrizing a more detailed model. For instance, we have not taken into account any difference in colonization prevalence caused by climate or demographic structure. Moreover, we were unable to control for differences in population structure, such as age, gender, and other institutions such as long-term care facilities as data on consumption and resistance at this level of detail was not available. Also, we used resistance to 3rd generation cephalosporins as a proxy for ESBL strains and assumed that these strains are the same across countries. An additional key limitation is the representativity of the resistance and consumption data used for this analysis: Resistance data were available only for bloodstream and spinal fluid infections. Moreover, consumption data were not complete for all years, and the collection process differs from country to country and is based on two different sources (reimbursement vs sale data) (29). However, we minimized the limitations associated with the consumption and resistance data by carefully restricting our analysis to countries with large numbers of isolates and consistent reporting over time. Moreover, the limitations of our approach are counterbalanced by the strengths of our data-based our modeling approach, which allows to provide a European perspective on the resistance problem in gram negative bacteria: using an epidemiological model, we could explain the variation and dynamics of antibiotic resistance in a key gram-negative pathogen at a European level and identify the drivers of its transmission. In particular, our work highlights the disproportionate role of antibiotic consumption in the hospital and of nosocomial transmission for resistance in gram negative bacteria. This indicates that infection control and antibiotic stewardship measures should play a major role in limiting resistance even at the national or regional level.

## Data Availability

All used data is publicly available.

https://gateway.euro.who.int/en/

https://www.ecdc.europa.eu/en/antimicrobial-resistance/surveillance-and-disease-data/data-ecdc

https://www.ecdc.europa.eu/en/antimicrobial-consumption/surveillance-and-disease-data/database

## Acknowledgments

We thank Katharina Kusejko, Huldrych Günthard, and Sebastian Bonhoeffer for useful discussions. As required by the ECDC, we confirm that “the views and opinions of the authors expressed herein do not necessarily state or reflect those of the ECDC. The accuracy of the authors’ statistical analysis and the findings they report are not the responsibility of ECDC. ECDC is not responsible for conclusions or opinions drawn from the data provided. ECDC is not responsible for the correctness of the data and for data management, data merging and data collation after provision of the data. ECDC shall not be held liable for improper or incorrect use of the data” (30). This study has been supported by the Swiss National Science Foundation (Grant no. BSSGI0_155851 to RDK).

